# A Phase I study targeting the APE1/Ref-1 redox signaling protein with APX3330: First clinical agent targeting APE1/Ref-1 in Cancer

**DOI:** 10.1101/2025.04.03.25325173

**Authors:** Mark R. Kelley, Jun Wan, Sheng Liu, Eyram Kpenu, Randall Wireman, Amber L Mosley, Hao Liu, Nehal J. Lakhani, Safi Shahda, Bert O’Neil, Mateusz Opyrchal, Richard A. Messmann

## Abstract

**Purpose:** APX3330 is an oral agent targeting the redox signaling activity of Ape1/Ref-1 (Ref-1), a key regulator of transcription factors involved in inflammation and tumorigenesis. APX3330 selectively inhibits Ref-1’s redox function without affecting its DNA repair role. This Phase 1, multicenter, open-label, dose-escalation study in advanced solid tumor was aimed at determining the recommended Phase 2 dose (RP2D) while assessing safety, pharmacokinetics, and biomarker evidence of target engagement. Clinical trial: NCT03375086.

**Patients and Methods:** Nineteen cancer patients were treated, with eight completing follow-up. Subjects received APX3330 orally twice daily in 21-day cycles, starting at 240 mg/day and escalating in 120 mg/day increments. Adverse event (AE) monitoring followed a 1 pt/cohort approach until a >G2 toxicity event, after which a 3+3 design was implemented. Treatment continued until disease progression, consent withdrawal, or intolerable toxicity. Antitumor activity was assessed using RECIST 1.1, and pharmacodynamic markers included serum Ref-1 levels and circulating tumor cells.

**Results:** Six subjects had stable disease for >4 cycles, with four remaining on study for 252– 421 days. No treatment-related serious adverse events occurred. One subject (720 mg cohort) withdrew due to Grade 3 maculopapular rash (dose-limiting toxicity). Laboratory assessments and ECGs showed no clinically significant abnormalities.

**Conclusions:** APX3330 demonstrated clinical benefit by stabilizing disease in ∼33% of subjects. Ref-1 target engagement was confirmed via biomarker analyses, with reduced serum Ref-1 and circulating tumor cells. The RP2D is 600 mg daily, with APX3330 showing a favorable safety profile and target-mediated effects.

## Introduction

Apurinic/apyrimidinic endonuclease/Redox effector factor 1 (APE1/Ref-1 or Ref-1) is a regulator of multiple transcriptional factors (TFs) through redox signaling that are critical to cancer survival and cross talks with many of the pathways implicated in resistance.(1–5) TFs targets of Ref-1 regulation include HIF-1α (hypoxia-inducible factor), NF-κB (nuclear factor κB), STAT3 (signal transducer and activator of transcription3), AP-1 (Fos/Jun, activator protein-1) and others. Ref-1 is both a redox (reduction-oxidation) signaling protein and a DNA repair enzyme (Figure. 1). (5–9) Ref-1’s redox function stimulates the DNA binding activity of numerous TFs by converting TFs from an oxidized to reduced form and enabling full TF activation. These TFs are often dysregulated in various solid and soft tumor cancers, and therefore blockade of Ref-1 redox activity can impede upon multiple cancer-related pathways.(7,10–13) Ref-1 plays a key role in a number of inflammatory disorders and tumorigenesis, including cancers of the colon, pancreas, skin, blood, as well as diabetic retinopathy, inflammatory bowel disorders and others.

**Figure 1.**
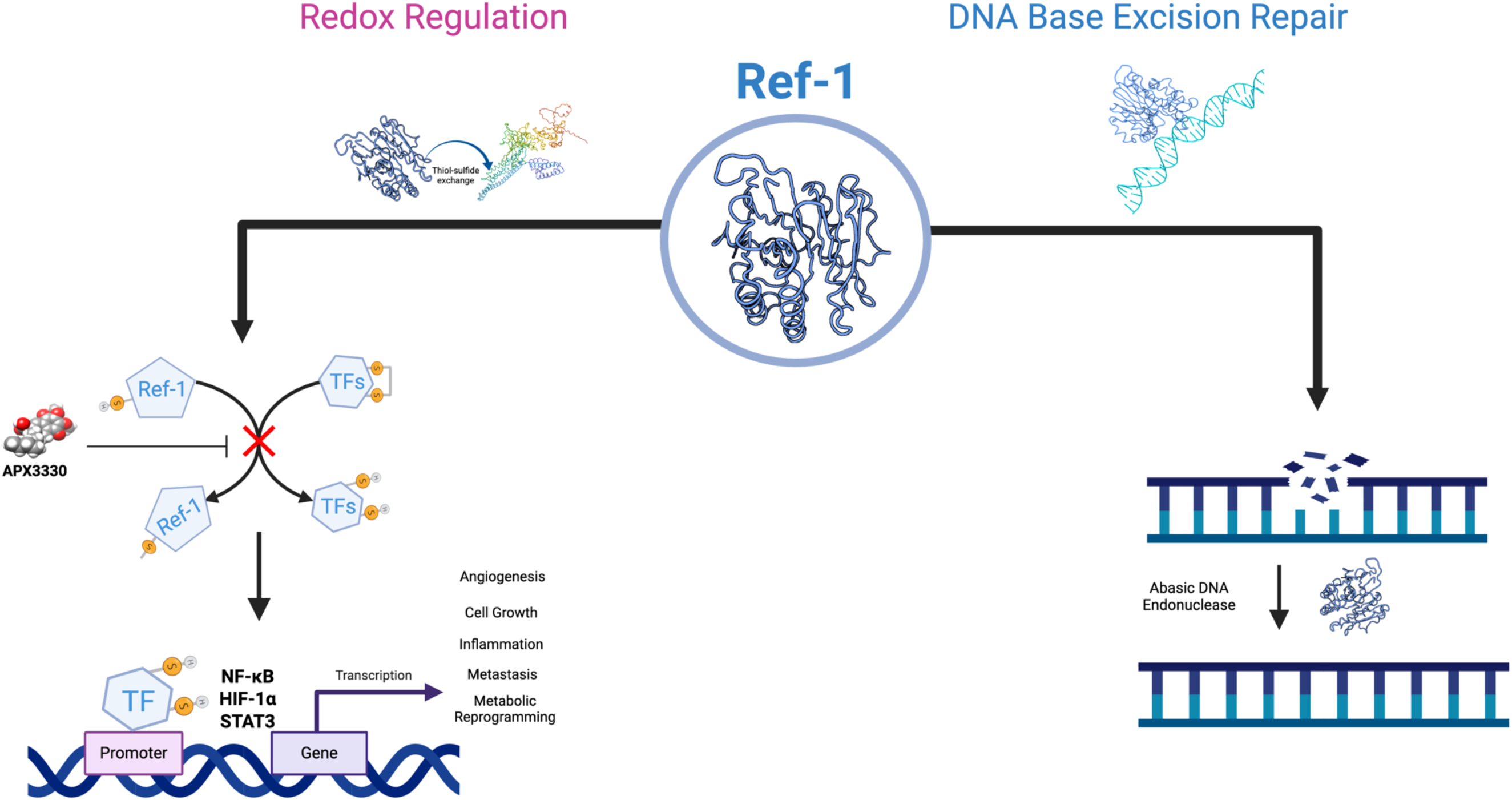
**Ref-1 target and specific inhibitor APX3330**. APX3330 blocks Transcription Factor (TF) activation in tumor cells while stimulating DNA Repair in normal cells, but not tumor cells.

Inhibition of protein-protein redox signaling activity is an innovative concept that is relatively unexplored as a target in cancer research, although it plays a role in many aspects of cancer signaling.(14) Ref-1 is an unusual redox factor in that it lacks an accessible nucleophilic Cys residue.(15) Our crystallographic studies suggest that Ref-1 is a well-folded, stable protein that must partially unfold for the redox active region to be exposed for activity and protein-protein interaction with the regulated TFs. Previous studies demonstrated that APX3330, our lead Ref-1 redox inhibitor and the subject of this clinical trial, produces a more open, unfolded Ref-1 protein that disables Ref-1’s activity.(16) This is a novel mechanism of action for protein inhibition (17). This makes Ref-1 a unique therapeutic target.

APX3330 is an orally administered agent targeting the Ref-1 protein. Administration of APX3330 results in clinical benefit to subjects with advanced solid tumors (e.g., endometrial, colorectal, prostate and melanoma cancer), as determined by mediated activity of Ref-1 targets and disease stabilization in approximately one-third of subjects for at least four 21-day treatment cycles. APX3330 is safe for chronic dosing up to 600 mg/d. Patient biopsy evaluation indicates APX3330-mediated effect upon cancer cells, including decrease in transcription factors regulated by the Ref-1 protein. Circulating tumor cell analysis indicates APX3330-mediated decrease in circulating tumor cells. All results consistently show that APX3330 mediates the activity of Ref-1 as expected based on pre-clinical data.

## Materials and Methods

### Patients, treatment schedules

This was an open-label, multicenter, dose-escalation study (NCT03375086), designed to determine the recommended Phase 2 dose (RP2D) of orally administered APX3330, as well as to evaluate the safety, tolerability, pharmacokinetic (PK) profile, and preliminary antitumor activity.

Up to approximately 30 eligible subjects with recurrent or advanced cancer (i.e., solid tumors) for whom standard therapy offered no curative potential, and who had adequate organ function, an Eastern Cooperative Oncology Group (ECOG) performance status (PS) of 0 to 2, and who were ≥ 21 days removed from therapeutic radiation or chemotherapy were planned for sequential enrollment into the dose-escalation phase.

### Trial design and end points

For all cohorts, oral APX3330 was administered twice daily (BID) on each day of a 21-day cycle. In the absence of Grade ≥3 adverse events (AEs) occurring during Cycle 1, subsequent cohorts were enrolled with at least 1 subject per cohort. Cohorts were to be expanded to a traditional 3+3 design in the event of any Grade ≥2 AE during Cycle 1 of a cohort for which there was no clear alternative explanation for causality (or after reaching any dose higher than APX3330 at 600-mg, whichever occurred first). In all cohorts, all subjects were required to complete the Cycle 1 dose-limiting toxicity (DLT) evaluation before dosing was initiated at the next-higher level. The RP2D was defined as a dose level that was well tolerated (i.e., at or below the maximum tolerated dose [MTD]) and for which the PK/pharmacodynamic data indicate adequate drug concentration and targeted molecular effect.

The dose escalation of APX3330 was planned to increase in 120mg increments between cohorts (i.e., total daily doses of 240-mg, 360-mg, 480-mg, 600-mg, 720-mg, 840-mg, 960-mg, etc.) in the absence of treatment-related toxicities. Subjects were permitted to continue APX3330 dosing until progressive disease (PD), intolerable toxicity, subject withdrawal, or termination of the study by the Sponsor.

The ability of APX3330 to inhibit its target was assessed by analysis of blood and tissue obtained from subjects, including the assessment of APX3330-mediated effects upon the apurinic/apyrimidinic endonuclease 1 (Ref-1) protein, and cellular functions controlled by Ref-1. The safety of APX3330 was assessed by the subject incidence of treatment-emergent adverse events (TEAEs), Common Terminology Criteria for Adverse Events (CTCAE; v4.03) grades at each dose level, dose adjustments, laboratory abnormalities, DLTs, serious adverse events (SAEs), and AEs leading to treatment discontinuation.

The study schema is provided in Figure 2.

**Figure 2.**
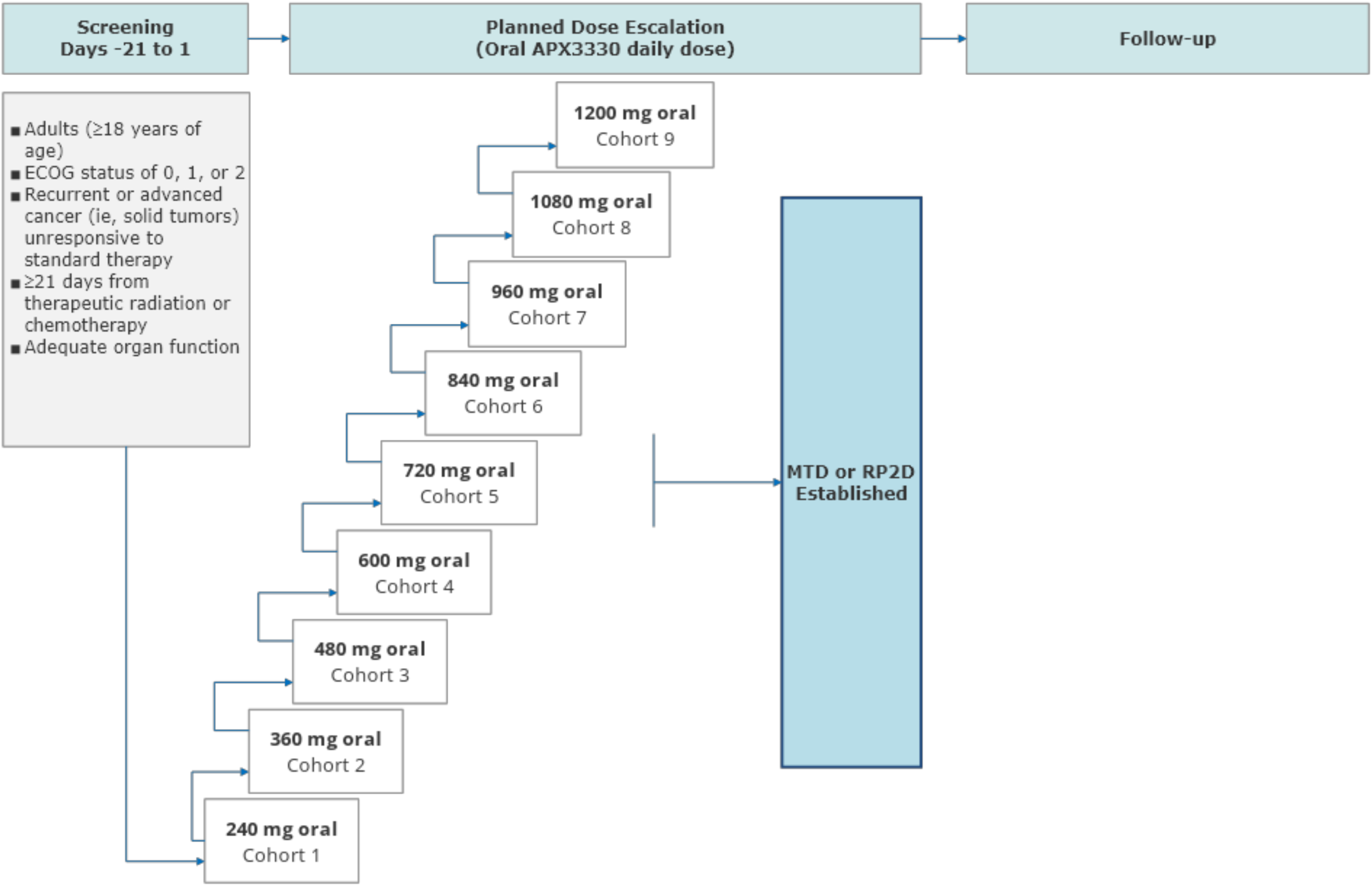
Study schema. Dose escalation proceeded until a RP2D was established. Cohort 1 begins with 1 subject. Additional cohorts continue enrolling 1 subject / cohort unitl the occurrence of either a) a > or = Grade 2 AE during Cycle 1 or b) dose escalation reaches 600 mg at which time the enrollment follows a 3+3 design.

### Proteomics

Quantitative proteomics analysis was performed using a TMT duplex Isobaric Mass Tagging Kit for comparison of pre-APX3330 treatment and post-APX3330 treatment tumor samples. In brief, tumor samples were solubilized in 8M Urea 100mM Tris-HCl pH 8.0. Following lysis by sonication, any remaining debris was removed by centrifugation for 30 minutes at 14,000 x g.

Proteins were then digested with Lys-C/Trypsin as we have previously described (PMID: 37562535). Following digestion, peptide samples were cleaned up using reversed phase columns (Waters) to remove the buffer components and then dried. Peptides were then resuspended in Triethylammonium bicarbonate and labeled using TMTduplex according to the manufacturer’s suggestions (Thermo Fisher). Following labeling, pre- and post-treatment samples were mixed at a 1:1 peptide ratio. The resulting peptide mixtures for each tumor pair were subjected to high pH reversed phase fractionation to obtain 6 fractions for mass spectrometry (MS) analysis. Samples were then analyzed by liquid chromatography-MS/MS on a Fusion Lumos MS as previously described (PMID: 31796630). Data were acquired with full MS acquisition at 60,000 resolution and MS/MS fragment analysis performed at a resolution of 50,000. MS/MS fragmentation was performed by higher-energy collision dissociation (HCD) with a collision energy of 36. Proteomics-based quantitation and protein database searching was performed with a human Uniprot FASTA database using the SEQUEST HT algorithm within Proteome Discoverer 2.2 (Thermo Fisher). Reporter ion quantitation was performed in PD 2.2 following sample normalization based on the total amount of protein detected for each reporter group. Samples were analyzed as individual pre- and post-treatment sets in addition to combination analyses of multiple tumor sets to determine specific changes that were tumor-specific or common amongst all analyzed groups.

### Circulating Tumor Cells

Circulating Tumor cells (CTCs) from 19 patients were processed by Epic Sciences (San Diego, CA) using their CTC chips (EpCAM+) technology. 35/37 (95%) samples passed technical quality control and were feasible for downstream analysis. 9/17 (53%) baseline samples (BL) had CTCs detected, while 9/18 (50%) on-treatment (OTx) draws had CTCs detected. 16 patients had BL and OTx samples that were further evaluated with the longitudinal analysis. Of these, 7/16 (44%), patients showed a reduction trend in all CTC populations (delta greater than or equal to 0 CTC/ml), respectively.

### Ref-1 serum levels

Blood samples were drawn pre-dose and six hours post-dose on D1, pre-dose D8 and D15 of cycle 1 for biomarker evaluation. The level of Ref-1 protein in whole blood was evaluated using a validated enzyme-linked immunosorbent assay (ELISA) assay (18,19). Ref-1 blood levels had previously shown a correlation with tumor aggressiveness (18,20). Comparison of Ref-1 protein levels with on-study levels were obtained to better understand the anti-tumor effect of APX3330 in patients with varying levels of Ref-1.

## Results

### Patient demographics

Nineteen subjects (13M, 6F) with median age of 67 years received therapy (Table 1). Dose (mg/d) escalation and number of patients treated (n) per each cohort proceeded as follows: 240- mg (1), 360 (4), 480 (2), 600 (6) and 720 (6). The median (range) number of doses administered was 84 (16‒695), and the median cumulative dose was 30,240 mg (5,760‒180,720 mg). The Safety Analysis Set was predominantly white (84.2%), male (68.4%), had a mean (SD) age of 67.8 (11.14) years, and a mean (SD) body mass index of 30.8 (7.65). All treated subjects had an ECOG PS of 0 (31.6%) or 1 (68.4%) at study entry, and the majority (68.4%) had nontarget lesions at baseline.

**Table 1.** Demographics of enrolled patients.

|  |  |
| --- | --- |
| Age<br>Mean (SD) | 67.8 (11.14) years |
| Gender<br>Female<br>Male | 6 Females (31.6%)<br>13 Males (68.4%) |
| Ethnicity<br>Not Hispanic/Latino<br>Hispanic/Latino<br>Not reported | 14 subjects (73.7%)<br>2 subjects (10.5%)<br>3 subjects (15.8%) |
| Race<br>White or Caucasian<br>Not reported | 16 (84.2%)<br>3 (15.8%) |
| Initial Starting Dose Levels (mg/d) – 19 subjects<sup>#</sup><br>240<br>360<br>480<br>600<br>720 | 1 subject<br>4 subjects<br>2 subjects<br>6 subjects<br>6 subjects* |
| Final Dose Levels (mg/d)<br>240<br>360<br>480<br>600<br>720 | 0<br>3<br>0<br>11<br>5 |
| Cancer Diagnosis | • 8 subjects with colorectal cancer<br>• 2 subjects with adenocarcinoma of the pancreas<br>• 1 subject with adenocarcinoma of the uterus<br>• 1 subject with gallbladder adenocarcinoma<br>• 1 subject with poorly differentiated carcinoma of the liver<br>• Metastatic adenocarcinoma<br>• 1 subject with melanoma<br>• 1 subject with adenocarcinoma of the prostate<br>• Papillary carcinoma |
|  | • High Grade carcinoma<br>• Poorly Differentiated carcinoma |

\*One subject at 720 mg dose level with maculopapular rash discontinued APX3330 treatment and withdrew from the study. Another subject at the 720 mg did develop a similar maculopapular rash and upon resolution of the rash, restarted at the 600 mg dose level and completed all study assessments. The two DLTs at the 720 mg dose level identified that level as being above the MTD. The 600 mg dose level was then expanded to enroll a sufficient number of subjects (>6) to identify 600 mg as the RP2D.
#Two subjects withdrew from the study in response to the occurrence of an AE (1 subject in the 600mg group with an unrelated small intestinal obstruction (an SAE), and 1 subject in the 720mg group with a related maculopapular rash). No subjects experienced a treatment-related SAE judged by the investigator to be related to APX3330.

At study entry, most (63.2%) subjects had an initial diagnosis of metastatic (stage IV) cancer (Supplemental Table 1). All subjects (100%) had received at least 1 prior chemotherapy regimen; the mean (SD) number of prior therapies was 6.3 (2.64). The majority of subjects discontinued treatment (17 of 19 total subjects [89.5%]); the most common reason for treatment discontinuation was disease progression (14 of 19 subjects).

### Dose Escalation Process

Dose escalation began with 1 subject per cohort, with the first evaluated APX3330 dose being 240 mg in Cohort 1 (Figure 2). For all cohorts, oral APX3330 was administered twice daily on each day of a 21-day cycle. In the absence of Grade ≥3 adverse events (AEs) occurring during Cycle 1, subsequent cohorts were enrolled with at least 1 subject per cohort. Cohorts were expanded via a traditional 3+3 design. The dose escalation of APX3330 was increased in 120- mg increments between cohorts (i.e., total daily doses of 240mg, 360mg, 480mg, 600mg, 720mg, 840mg, 960mg, etc.) in the absence of treatment-related toxicities (Supplemental Figure 1).

### Safety

APX3330 was well tolerated at dose levels from 240-600 mg/d (Supplemental Table 2). The most frequent treatment-related adverse events (all grades) included G1 nausea (16%) and fatigue (16%). Of the 19 subjects included in the principal analysis of safety, 15 (78.9%) had ≥1 TEAE (Supplemental Table 2). The incidence of TEAEs was not dose-related (120-mg: 1 subject [100%]; 360-mg: 3 subjects [75.0%]; 480mg: 2 subjects [100%]; 600-mg: 5 subjects [83.3%]; 720-mg: 4 subjects [66.7%]). Of the 15 patients with TEAEs, 9 (47.4%) experienced events considered by the investigators to be related to APX3330.

Two subjects in the 720mg group experienced DLTs of treatment-related Grade 3 maculopapular rash during their first cycle of treatment. For 1 of these subjects, the maculopapular rash resulted in discontinuation of APX3330 and withdrawal from the study. For the second subject, the 720mg dose of APX3330 was interrupted until resolution of the TEAE, whereupon treatment was restarted at a the next-lower dose (i.e., 600-mg); this subject completed all study assessments at the 600mg APX3330 dose without recurrence of the rash. Both events were resolved.

Two subjects withdrew from the study in response to the occurrence of an AE (1 subject in the 600mg group with an unrelated small intestinal obstruction (an SAE), and 1 subject in the 720mg group due to a related maculopapular rash). No subjects experienced an SAE judged by the investigator to be related to APX3330. The 2 DLTs at the 720mg dose level identified that level as being above the MTD. The 600mg dose cohort was then expanded to enroll an additional 3 patients and 600 mg was identified as the MTD.

### Clinical Laboratory Evaluations

Changes from baseline in hematology, chemistry, and urinalysis laboratory results were not clinically meaningful. There were no laboratory results reported as AEs that were considered by the investigators to be related to APX3330. There were no clinically significant 12-lead electrocardiogram (ECG) abnormalities reported, and vital signs changes from baseline were generally unremarkable.

### Efficacy

Of the 19 subjects treated with oral APX3330 at daily doses ranging from 240 to 720-mg, 6 (31.6%) had stable disease or better for more than four 21day treatment cycles, and one patient (10.5%) had confirmed PR for a clinical benefit rate of 42.1%. (Figure 3A). Notably, 4 subjects remained on study for a duration of 252 to 421 days with stable disease or better. Including:

- One subject with colorectal cancer who remained on study for 357 days and had a PR
- One subject with endometrial cancer who remained on study for 421 days and had stable disease
- One subject with melanoma who remained on study for 337 days and had stable disease
- One subject with prostate cancer who remained on study for 252 days and had stable disease

**Figure 3.**
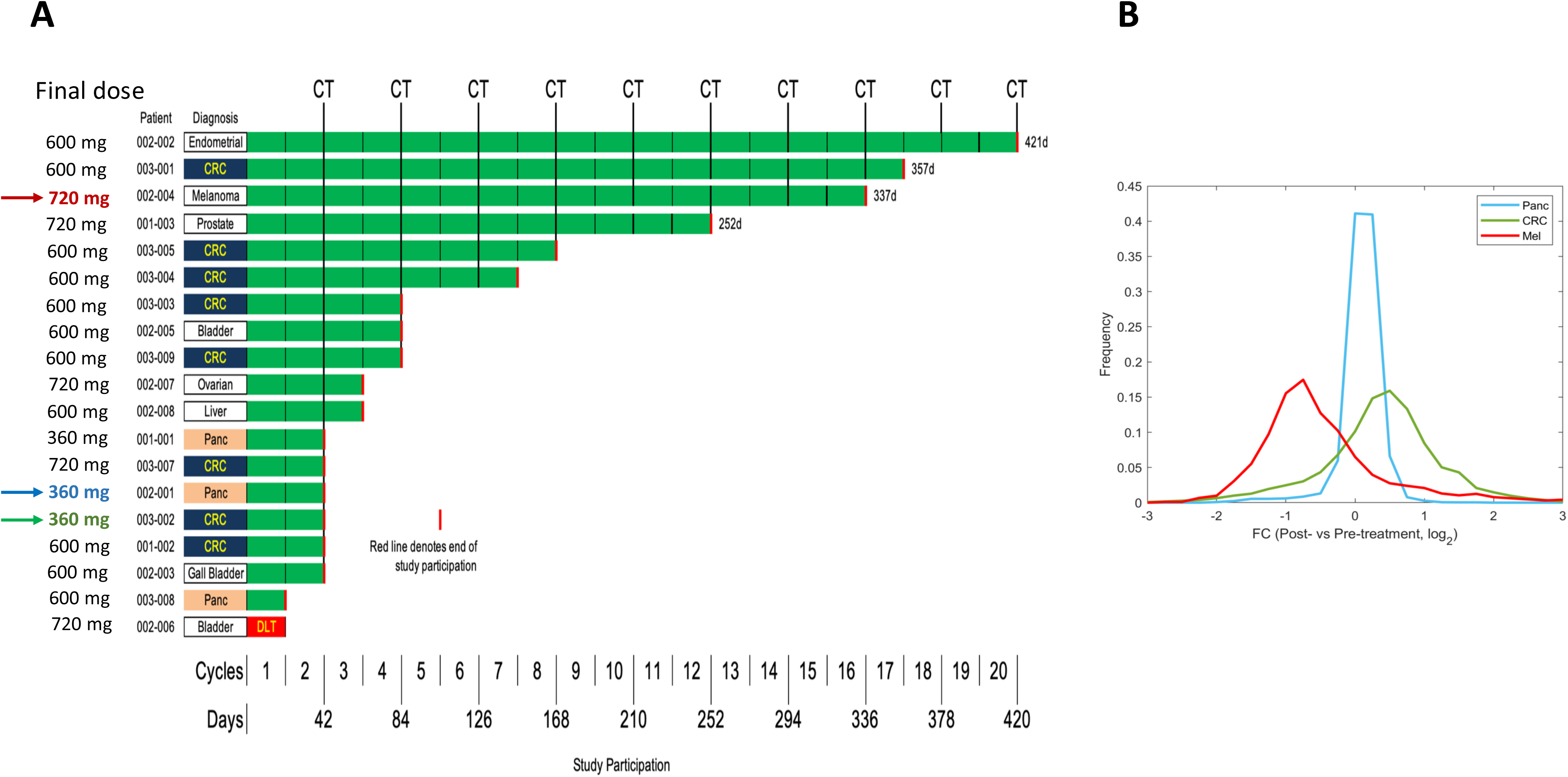
Swimlane plot of 19 patients treated with APX3330 in a dose ascending trial. **A)** CRC = colorectal cancer; CT = chemotherapy; DLT = dose-limiting toxicity; Panc = pancreatic cancer CRC patient 003-001 had a partial response (PR) and marked in red. B) Proteomics data analysis. Distribution of fold changes (FCs, in the scale of log2) of protein expression levels post-treatment vs pre-treatment. Melanoma patient (red) has more significant response to the drug than other two patients analyzed (Pancreatic, blue and Colon, green).

Serum levels of Ref-1 are elevated in patients with minimal clinical benefit from APX3330 and reduced in patients with documented clinical benefit Whole blood analysis of Ref-1 protein levels (as determined by ELISA assay) were evaluated at baseline and on-treatment. Ref-1 levels were compared between subjects with PD and those with stable disease (defined as those who remained on treatment for >4 cycles) or better, the mean maximum Ref-1 level was significantly (p=0.028) higher among subjects with PD (Supplemental Figure 1A) showing that orally administered APX3330 mediated the activity of the APE1/Ref-1 target.

### Circulating Tumor Cell Analysis

Cumulatively, 37 samples from 19 patients were processed for CTCs by Epic Sciences. 35/37 (95%) samples passed technical quality control and were feasible for downstream analysis.

9/17 (53%) baseline samples (BL) had CTCs detected, while 9/18 (50%) on-treatment (OTx) draws had CTCs detected. 16 patients had BL and OTx samples that were further evaluated with the longitudinal analysis. Of these, 7/16 (44%), 6/16 (38%), and 3/16 (19%) patients showed a reduction trend, an increase trend, and no change in all CTC populations (delta greater than or equal to 0 CTC/ml), respectively (Supplemental Figure 1B). Of the 7 patients with a reduced trend of CTCs, five of the patients were on APX3330 for 3 cycles or greater (3, 3, 4, 12, and 20) while the other two patients were on for 1 and 2 cycles, respectively.

### Confirmed target engagement: Proteins altered downstream of Ref-1 regulated transcription factors

Samples were collected from three patients with Pancreatic cancer (Panc), Colon cancer (CRC), and Melanoma (Mel), respectively. Using the proteomics data achieved from these samples, we preformed paired biopsy analysis between pre-treatment and on-treatment samples. Differences were observed (Figure 3B and supplemental Figure 2) in terms of fold change (FC) distributions of protein expressions for these three cancer patients. The most proteins for Mel tended to be down-regulated on-treatment which correlated with that patient having the longest period of non-tumor growth. All patients analyzed had a significantly different pattern of protein expression from the untreated patient samples (Supplemental Figure 2).

We defined differentially expressed proteins (DEPs) if the amplitudes of their linear fold changes by comparing pre-treatment and on-treatment tumor biopsies were larger than 1.5 or less than 2/3 (Supplemental Figure 2). FCs of 2580 DEPs obtained from any cancer patient were shown in Figure 4A, where some DEPs might be detected in one cancer but not in others (white bars in Figure 4A). Among them, only 225 proteins presented at least 1.5-FC for Panc, from which 64.9% were repressed on treatment. However, many more proteins showed larger FCs for CRC (n = 1,828 with 23.2% down-regulation) and Mel (n = 1,278 with 83.6% down-regulation).

**Figure 4.**
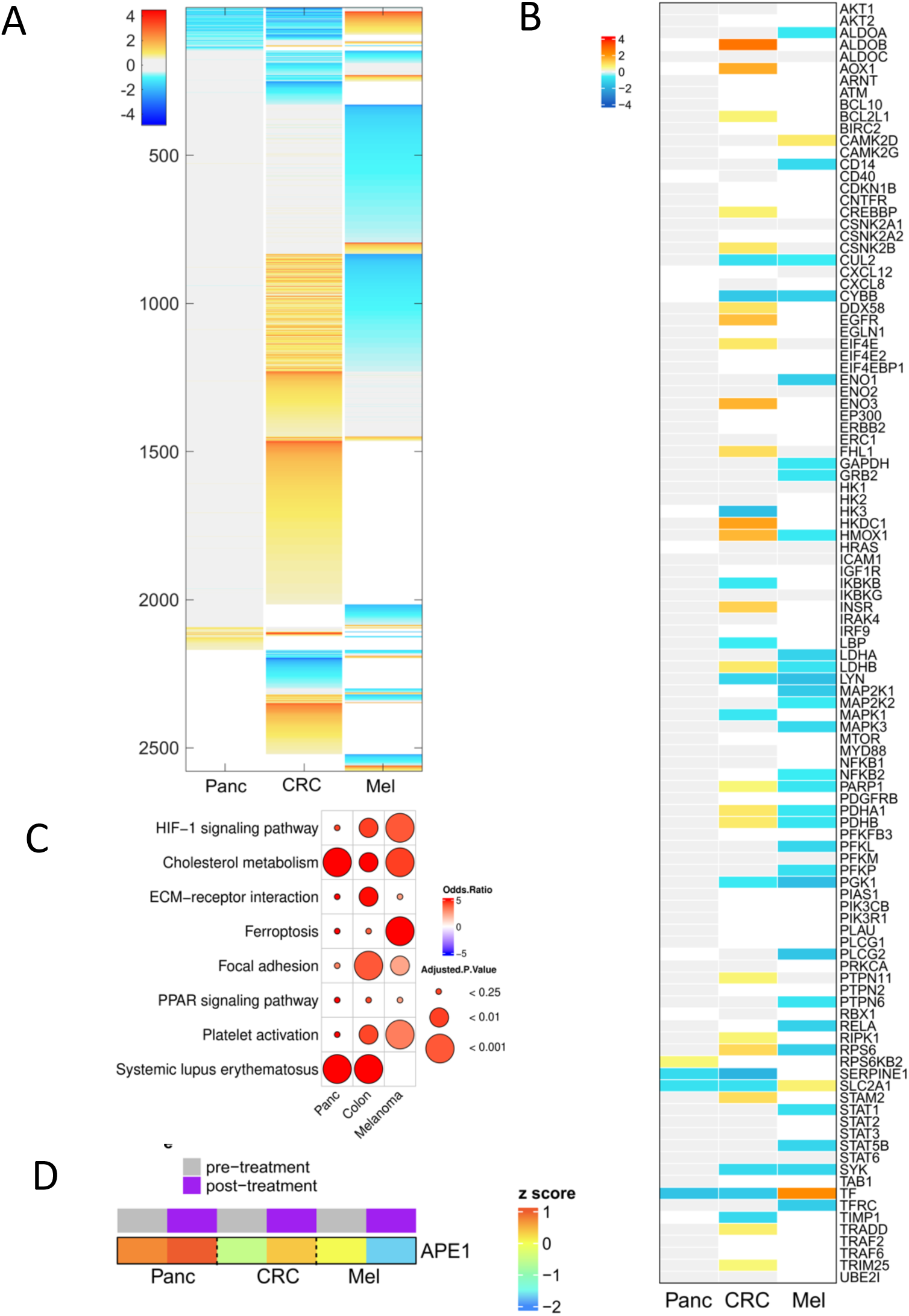
A. Heatmap of DEPs identified in at least one cancer/patient sample for Panc, Colon, and Melanoma (1^st^ sample shortly after treatment and 2^nd^ sample about one year after treatment). FCs with less than 1.5-fold and greater than 2/3-fold were marked by the color of gray. The white represents proteins not detected. **B**. DEPs with corresponding FCs involved in three pathways, HIF1a, NFkB and STAT3. **C**. Pathways significantly enriched in DEPs. **D**. APE1 protein expression pre- and post-treatment for three cancer patients.

Interestingly, a few proteins, TNC (Tenascin), RHOA (Transforming protein RhoA), and TXNRD1 (Thioredoxin reductase 1, cytoplasmic), exhibited same direction changes, all of which were downregulated when comparing APX3330 treatment to pre-treatment. Out of 537 DEPs identified in Mel biopsy that also showed more than 1.5-FC for either Panc or CRC, 85.3% showed opposition expression change direction between Mel and Panc/CRC. Particularly for genes associated with HIF1α/NF-κB/STAT3 pathways (Figure 4B), some were uniquely downregulated in melanoma patient only, such as GRB2 (growth factor receptor bound protein 2), LDHA (lactate dehydrogenase A), and NF-κB2 (nuclear factor kappa-B subunit 2), or upregulation in CRC, e.g., HMOX1 (heme oxygenase 1), PARP1 (Poly ADP-Ribose) Polymerase 1), and RPS6 (ribosomal protein S6). Taken together, these results suggest diverse target proteins in the different solid tumors impacted when treated with APX3330.

The enrichment analysis (Figure 4C) revealed that some KEGG pathways were significantly (adjusted p-value less than 0.05) enriched in down-regulated DEPs on treatment for all three cancer patients, including HIF-1α signaling pathway, inflammation-stimulated cholesterol metabolism, ECM-receptor interaction, ferroptosis, focal adhesion, PPAR signaling pathway, platelet activation, and autoimmune-related systemic lupus erythematosus. The HIF-1α signaling pathway was remarkably over-represented in DEPs from all three patients. However, the targeted DEPs can be the same or very different (Figure 4B). For example, several mitogen-activated protein kinases, MAP2K1/2 and MAPK3 were repressed for Mel patient, but not for Panc or CRC patient (Figure 4C). Proteins CUL2 (Cullin 2), CYBB (Cytochrome B-245 Beta Chain), and PGK1 (Phosphoglycerate Kinase 1) were observed to be downregulated for both Mel as well as CRC. Another two proteins SLC2A1 (Solute Carrier Family 2 Member 1) and TF (TF protein) were repressed for pancreatic cancer and colon cancer but was activated for melanoma. This suggests that APX3330 might be able to control the same signaling pathway by targeting divergent protein components in the pathway and this response reflects the impact on the tumor growth. It is worth noting that platelet activation was associated with the NF-κB pathway (21). While the NF-κB pathway itself was not quite significant in down-regulated DEPs by APX3330, we still observed several repressed DEPs involved in NF-κB or in JAK-STAT signaling pathway (Figure 4B), such as Lipopolysaccharide Binding Protein (LBP), LYN (LYN Proto-Oncogene, Src Family Tyrosine Kinase), NF-κB2 (nuclear factor kappa-B subunit 2), PLCG2 (Phospholipase C Gamma 2), PTPN6 (Protein Tyrosine Phosphatase Non-Receptor Type 6), RELA (RELA Proto-Oncogene, NF-κB Subunit), STAT1/5B (Signal Transducer And Activator Of Transcription 1/5B), and SYK (Spleen Tyrosine Kinase).

Finally, the level of APE1 protein was found to be greatly reduced in the Mel patient who was on the APX3330 treatment for a significantly longer time compared to the Panc and CRC patients who did not respond as well (Figure 4D). APX3330 does not normally cause downregulation of the protein expression of APE1, but this data supports data observed in other studies (22) suggesting that with the slowing of tumor growth, less APE1 is required to activate the TFs such that a negative feedback loop occurs.

## Discussion

The results presented here represent the first-in human clinical trial for APX3330 targeting the Ref-1 protein in cancer. Subsequent to this trial completion and positive data obtained as an oral agent, APX3330 underwent a phase II trial in diabetic retinopathy and diabetic macular edema and reported elsewhere (23). Ref-1 is a redox signaling protein, and its role in the regulation of numerous transcription factors (TFs) such as HIF-1α, NF-κB, STAT3, has been shown from numerous preclinical studies (24–32) (Figure 1). Regulation of HIF-1α by Ref-1 along with these other TFs contributes to the metabolic rewiring and gene expression changes that are observed in many cancers.(33–36) Ref-1 is both a redox (reduction-oxidation) signaling protein and a DNA repair enzyme. It contributes to the repair of DNA damage caused by alkylators, internal base loss and AP site formation, and oxidizing cellular DNA damaging conditions as well as external oxidizing chemo agents within the DNA base excision repair (BER) pathway.(5–9) Ref-1 is also a redox factor that stimulates the DNA binding activity of numerous TFs, by converting TFs from an oxidized to reduced form and enabling full TF activation (Figure 1). These TFs are often dysregulated in cancer, and therefore blockade of Ref-1 redox activity can impede upon many cancer-related pathways.(7,10–13) In this phase I clinical trial (NCT03375086), subjects were enrolled in the dose-escalation phase to receive oral APX3330, with the first evaluated dose being 240-mg in Cohort 1. Patients received APX3330 continuously on twice daily regimen in 21-day cycles starting at 240-mg/day and escalating in 120-mg/day increment (Figure 2). AE evaluation included 1 pt/cohort until the occurrence of ≥ G2 toxicity at which time the study proceeded in a 3+3 design. Subjects were permitted to continue treatment until progression of disease (PD), withdrawal of consent, or intolerable toxicity. Safety was assessed throughout the study; antitumor activity was based on radiological imaging at the end of every second cycle using standard RECIST 1.1 criteria; pharmacodynamic assessments included blood sampling to assess serum Ref-1 protein levels and circulating tumor cell counts. Primary and secondary objectives included determining the recommended phase 2 dose (RP2D), the safety and PK/PD profiles of APX3330 and reporting any RECIST anti-tumor activity.

In total, 19 subjects were enrolled and treated in this study, constituting the Safety Analysis Set. In total, eight subjects completed the 30-day follow-up visit. While receiving oral BID administration of APX3330, six (of 19) subjects with advanced tumors had disease stabilization for more than four 21day treatment cycles, with four of these subjects remaining on study for a duration of 252 to 421 days with stable disease. There were no AEs leading to death, and there were no treatment-related serious adverse events (SAEs). The most frequent treatment-related adverse events (all grades) included G1 nausea (16%) and fatigue (16%). A G3 rash occurred in two subjects at the 720 mg level defining 600 mg/d as the RP2D for further development. One subject (720mg group) withdrew from the study in response to a treatment related AE of Grade 3 maculopapular rash (a dose-limiting toxicity [DLT]). Changes from baseline in hematology, chemistry, and urinalysis laboratory results were not clinically meaningful. There were no laboratory results reported as AEs that were considered by the investigator to be related to APX3330. There were no clinically significant 12-lead ECG abnormalities reported, and vital signs changes from baseline were generally unremarkable. The protein expression changes highlighted distinct effects of APX3330 on Panc, Col, and Mel patients.

The most significant effect was observed in the Mel patient, where targeted proteins were predominantly down-regulated upon the treatment, especially those involved in the NF-κB and JAK-STAT signaling pathway. Several KEGG pathways were significantly over-represented in down-regulated DEPs on treatment across three cancer patients, including HIF-1α signaling pathway, inflammation-stimulated cholesterol metabolism, ECM-receptor interaction, ferroptosis, focal adhesion, PPAR signaling pathway, platelet activation, and autoimmune-related systemic lupus erythematosus. Notably, the HIF-1α signaling pathway was remarkably enriched for all three patients, suggesting APX3330 may modulate this pathway by targeting different protein components, ultimately influencing tumor growth. We also detected an overall reduction in APE1 levels in the treated patients as well as decreased circulating tumor cells, although these findings will need to be further evaluated as biomarkers of the APX3330 in future trials due to the small sample size of this study.

In summary, all study objectives were completed for the phase I trial of APX3330. Administration of APX3330 resulted in clinical benefit to subjects with advanced solid tumors, as determined by mediated activity of Ref-1 targets and disease stabilization in approximately one third of subjects for at least four 21day treatment cycles. Administration of APX3330 at daily doses up to and including 600-mg is safe and well tolerated and patients were on the drug for extended periods of time, up to and over a year, and it provided clinical benefit to patients in a variety of tumor types (e.g., endometrial, colorectal, prostate and melanoma cancer). Data from the recently completed phase II trial in DR/DME also demonstrated a strong safety profile for patients taking 600 mg/day for 6 months (23). Furthermore, all results consistently show that APX3330 mediates activity of Ref-1 target as expected based on pre-clinical data. Predicted PK was observed as expected and previously published (37,38). Patient biopsy evaluation indicates APX3330-mediated effect upon cancer cells, including decrease in transcription factor activity regulated by the APE1 protein such as HIF-1α, NF-κB and STAT3. Circulating tumor cell analysis indicates APX3330-mediated decrease in tumor cells. All results consistently show that APX3330 mediates activity of APE1/Ref-1 target as expected.

## Data Availability

The data supporting this study are not publicly available due to proprietary restrictions but can be obtained from the corresponding author upon reasonable request.

## Acknowledgements

We want to thank Lincy Chu and Amanda Anderson at Epic Sciences for performing the circulating tumor cell analysis. We would also like to thank Aruna Wijeratne for help on running the proteomics samples in the IUSCCC Proteomics Core.

## Ethics statement

This study was conducted with approval by the Institutional Review Board (IRB) (Approval Number: 00003544 (07-16-2016). All participants were provided written informed consent before enrollment in the study. The trial was registered at ClinicalTrials.gov under the identifier NCT03375086.

---

**Supplemental** Figure 1**: Ref-1 Serum Levels and CTC in patients treated with APX3330.** A. Ref-1 serum levels in subjects with progressive disease versus those with stable disease or better. Ref-1 serum levels were determined using a standard ELISA assay. Statistical comparisons were done between two groups (patients with SD vs PD) using two-sample t-test. p-value 0.028 and is statistically significant. SD patients are defined as those on treatment past 4 cycles. B. APX3330 reduced circulating tumor cells (CTC) in patients. 16 out of 19 patients (84%) had baseline and Otx samples. 7 out of 16 patients (44%) of patients showed a reduction in all CTC populations.

**Supplemental** Figure 2**: Protein expression levels (log2 scale) for pre-treatment (x axis) and post-treatment (y axis).** The red and blue dots represent differentially expressed proteins (DEPs), either up-regulated (red, greater than 1.5-fold) or down- regulated (blue, less than 2/3-fold) after the treatment.

