## Supplementary figures and images for "A Phase I study targeting the APE1/Ref-1 redox signaling protein with APX3330: First clinical agent targeting APE1/Ref-1 in Cancer"

### Supplemental figures

#### Supplemental Figure 1

A

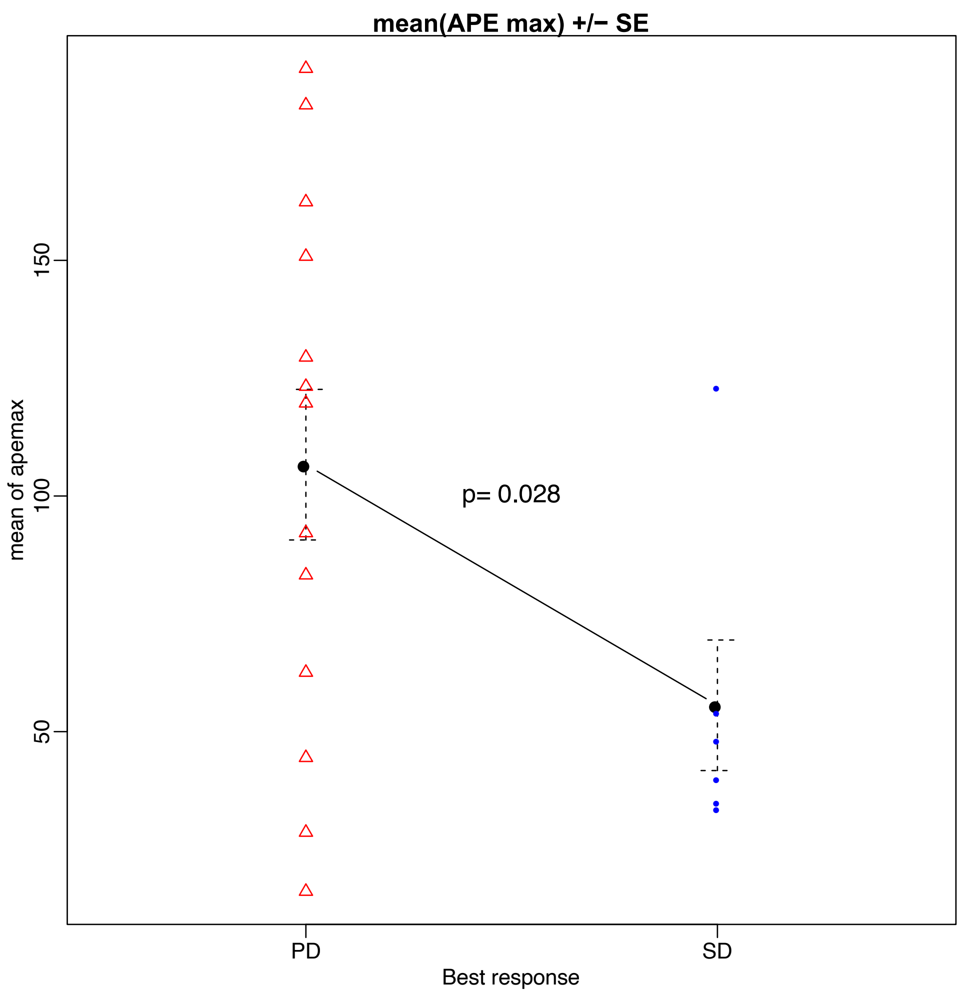


B

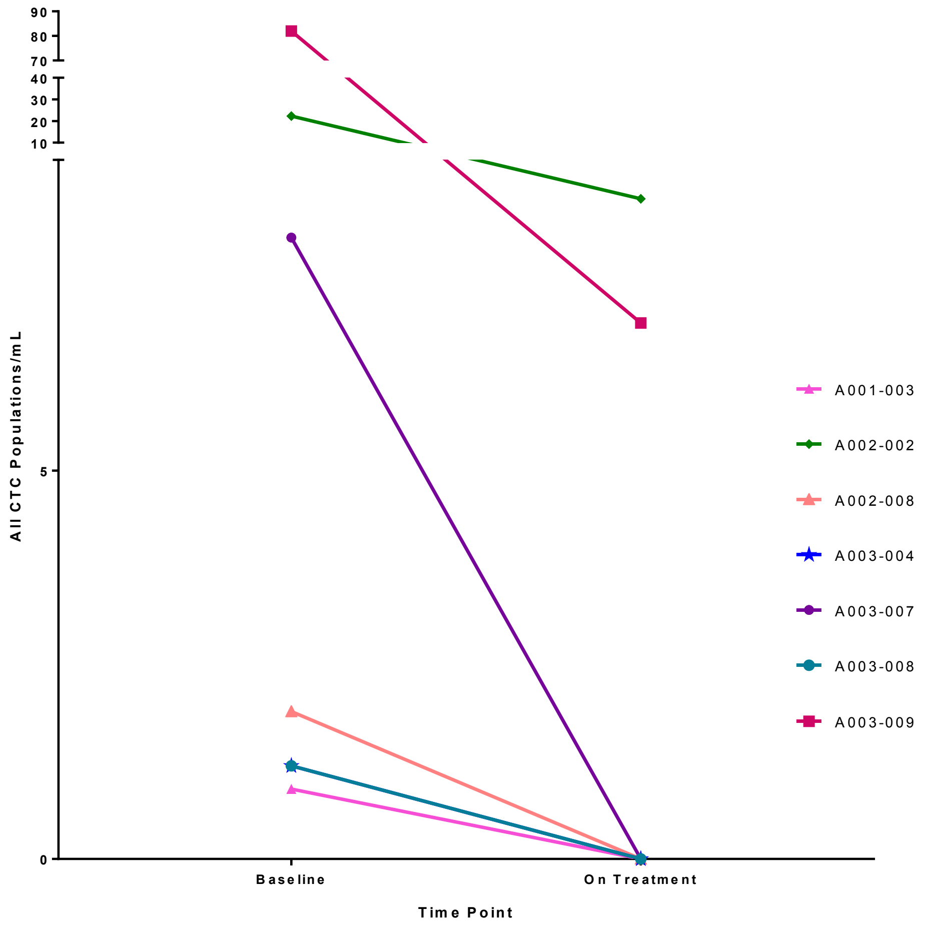

#### Supplemental Figure 2

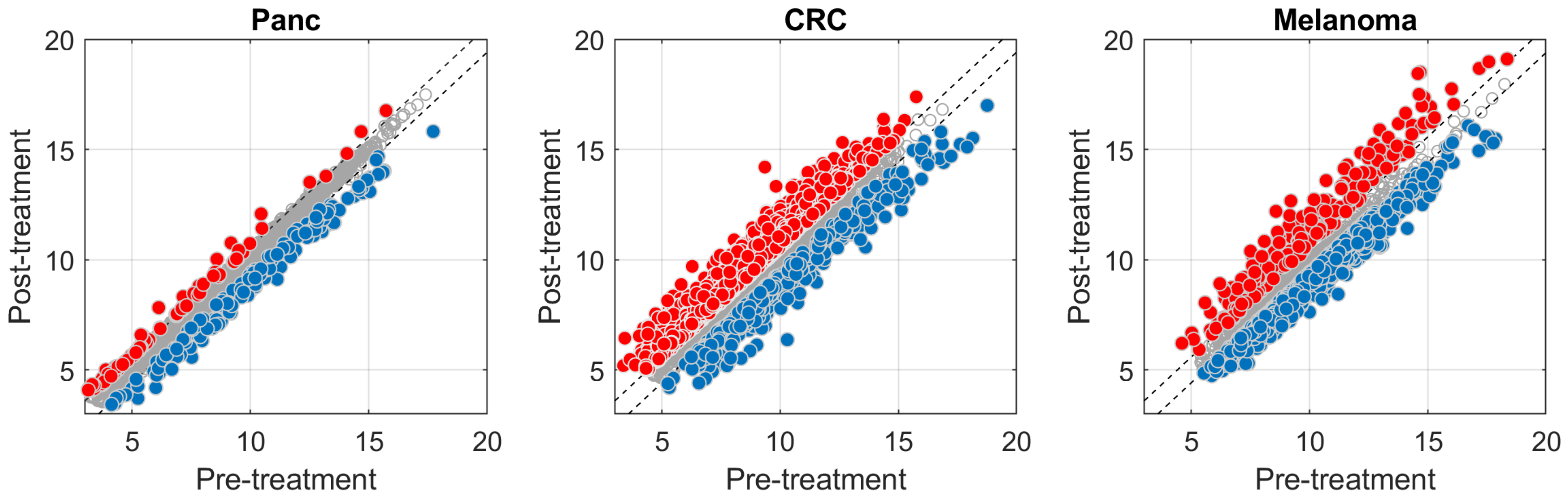
