## Supplemental Table 2 for "A Phase I study targeting the APE1/Ref-1 redox signaling protein with APX3330: First clinical agent targeting APE1/Ref-1 in Cancer"

### Phase I trial and Cancer Diagnosis

| | 240 mg N=1 | 360 mg N=4 | 480 mg N=2 | 600 mg N=6 | 720 mg N=6 | All subjects N=19 |
|---|---|---|---|---|---|---|
| Primary Diagnosis [n (%)] | | | | | | |
| Adenocarcinoma of Pancreatic Head | 0 | 1 (25.0%) | 0 | 0 | 0 | 1 (5.3%) |
| Adenocarcinoma of the colon | 0 | 0 | 0 | 1 (16.7%) | 0 | 1 (5.3%) |
| Adenocarcinoma of the prostate | 0 | 0 | 0 | 0 | 1 (16.7%) | 1 (5.3%) |
| Adenocarcinoma of the uterus | 0 | 0 | 1 (50.0%) | 0 | 0 | 1 (5.3%) |
| Colon Adenocarcinoma | 0 | 0 | 0 | 0 | 1 (16.7%) | 1 (5.3%) |
| Colon Cancer-adenocarcinoma | 0 | 0 | 0 | 1 (16.7%) | 0 | 1 (5.3%) |
| Colorectal | 0 | 0 | 0 | 1 (16.7%) | 0 | 1 (5.3%) |
| Gallbladder adenocarcinoma | 0 | 0 | 0 | 1 (16.7%) | 0 | 1 (5.3%) |
| High Grade Carcinoma | 0 | 0 | 0 | 0 | 1 (16.7%) | 1 (5.3%) |
| Intraoperative mucosal well-differentiated adenocarcinoma Colon. | 0 | 1 (25.0%) | 0 | 0 | 0 | 1 (5.3%) |
| Melanoma of the left posterior leg | 0 | 0 | 0 | 0 | 1 (16.7%) | 1 (5.3%) |

| | | | | | | |
|---|---|---|---|---|---|---|
| Metastatic adenocarcinoma | 0 | 0 | 0 | 1 (16.7%) | 0 (0.0%) | 1 (5.3%) |
| Papillary Carcinoma | 0 | 0 | 0 | 0 | 1 (16.7%) | 1 (5.3%) |
| Poorly Differentiated Carcinoma | 0 | 0 | 0 | 0 | 1 (16.7%) | 1 (5.3%) |
| Poorly differentiated carcinoma of the liver | 0 | 0 | 0 | 1 (16.7%) | 0 | 1 (5.3%) |
| Rectal Adenocarcinoma | 0 | 1 (25.0%) | 0 | 0 | 0 | 1 (5.3%) |
| Rectal Moderately differentiated adenocarcinoma | 1 (100.0%) | 0 | 0 | 0 | 0 | 1 (5.3%) |
| Well differentiated adenocarcinoma of the pancreas | 0 | 1 (25.0%) | 0 | 0 | 0 | 1 (5.3%) |
| Invasive low grade adenocarcinoma Rectal Cancer | 0 | 0 | 1 (50.0%) | 0 | 0 | 1 (5.3%) |
| Time Since Initial Diagnosis of Cancer[1,2] | | | | | | |
| n | 1 | 2 | 2 | 6 | 5 | 16 |
| Mean (SD) | 3.0 (--) | 1.5 (0.71) | 3.5 (3.54) | 2.8 (3.19) | 4.2 (3.11) | 3.2 (2.76) |
| Min, Max | 3.0, 3.0 | 1.0, 2.0 | 1.0, 6.0 | 0.0, 9.0` | 1.0, 8.0 | 0.0, 9.0 |
| Median | 3.0 | 1.5 | 3.5 | 2.0 | 3.0 | 2.0 |
| Stage at Initial Diagnosis [n (%)] | | | | | | |
| 0 | 0 | 0 | 0 | 0 | 0 | 0 |
| I | 0 | 1 (25.0%) | 0 | 0 | 0 | 1 (5.3%) |
| II | 0 | 1 (25.0%) | 1 (50.0%) | 1 (16.7%) | 0 | 3 (15.8%) |
| III | 0 | 0 | 0 | 1 (16.7%) | 2 (33.3%) | 3 (15.8%) |
| IV | 1 (100.0%) | 2 (50.0%) | 1 (50.0%) | 4 (66.7%) | 4 (66.7%) | 12 (63.2%) |
| Number of Previous Therapies[3] | | | | | | |
| n | 1 | 4 | 2 | 6 | 6 | 19 |
| Mean (SD) | 5.0 (--) | 6.5 (2.08) | 5.0 (2.83) | 4.8 (1.72) | 8.2 (3.19) | 6.3 (2.64) |
| Min, Max | 5.0, 5.0 | 4.0, 9.0 | 3.0, 7.0 | 2.0, 7.0 | 5.0, 12.0 | 2.0, 12.0 |

| | | | | | | |
|---|---|---|---|---|---|---|
| Median | 5.0 | 6.5 | 5.0 | 5.0 | 8.0 | 6.0 |
| Patients With As Least One Prior Chemotherapy Regimen[4] [n (%)] | 1 (100.0%) | 4 (100.0%) | 2 (100.0%) | 6 (100.0%) | 6 (100.0%) | 19 (100.0%) |
| Prior Best Response for Chemotherapy[5] [n (%)] | | | | | | |
| Complete Response | 0 | 0 | 0 | 0 | 1 (4.2%) | 1 (1.8%) |
| Partial Response | 0 | 1 (9.1%) | 0 | 0 | 1 (4.2%) | 2 (3.5%) |
| Stable Disease | 3 (75.0%) | 4 (36.4%) | 4 (80.0%) | 3 (23.1%) | 7 (29.2%) | 21 (36.8%) |
| Disease Progression | 0 | 3 (27.3%) | 0 | 7 (53.8%) | 2 (8.3%) | 12 (21.1%) |
| Not Applicable | 1 (25.0%) | 0 | 0 | 0 | 1 (4.2%) | 2 (3.5%) |
| Unknown | 0 | 3 (27.3%) | 1 (20.0%) | 3 (23.1%) | 12 (50.0%) | 19 (33.3%) |
| Time to Progressive Disease[6 7] | | | | | | |
| n | 2 | 4 | 1 | 5 | 6 | 18 |
| Mean (SD) | 0.5 (0.71) | 0.3 (0.50) | 0.0 (--) | 0.0 (0.00) | 0.5 (0.55) | 0.3 (0.46) |
| Min, Max | 0.0, 1.0 | 0.0, 1.0 | 0.0, 0.0 | 0.0, 0.0 | 0.0, 1.0 | 0.0, 1.0 |
| Median | 0.5 | 0.0 | 0.0 | 0.0 | 0.5 | 0.0 |

1 'Time Since Initial Diagnosis of Cancer' is calculated from the formula: (the date of signing the informed consent - the date of initial cancer diagnosis + 1) / 365.

2 'Time Since Initial Diagnosis of Cancer' was not calculated for patients 22497, 22619 and 115034 due to an incomplete date of initial cancer diagnosis.

3 Includes prior chemotherapy, radiation and cancer-related surgeries.

4 The next two sections will summarize data only for patients who received at least one prior chemotherapy regimen.

5 The best response over all chemotherapy regimens is reported.

6 'Time to Progressive Disease' was calculated from the formula: (prior disease progression image date - prior chemotherapy start date + 1) / 365.

7 Due to complete or partial missing data in prior chemotherapy start date and prior disease progression image date only 18 cases were reported. Program: TREF05.sas Data extract date: Table Generation: 02DEC2021 11:55
